# Clinical Appearance of Malaria and its Epidemiological Burden in Mangalore, India in 2002 to 2022

**DOI:** 10.1101/2025.08.12.25333441

**Authors:** Benudhar Mukhi, Chandan Kumar, Mahesh Chandra Sahu, Prince Kumar, D Pradeep, Anupkumar R. Anvikar, Susanta Kumar Ghosh

## Abstract

**Background:** Incomplete antimalarial medication and insufficient patient follow-up, particularly among migrant labourers and destitute inpatients, pose significant challenges to malaria treatment and hinder elimination efforts in India. Patients who escape from hospitals during intermittent treatment or fail to receive medication despite tested positive for malaria parasites (MP) contribute to transmission, antimalarial resistance, genetic mutations, recurrent malaria, early treatment failure (ETF) and late treatment failure (LTF).

**Methods:** A clinical case study was conducted at Wenlock District Government Hospital, Mangalore, India, from 2002 to 2022. Suspected malaria cases were confirmed using symptomatic clinical features and microscopic examination of peripheral blood smears. Clinical parameters were monitored and a second smear was taken on day 7 to assess drug response and treatment efficacy. AI-driven tools analysed 21 years of malaria cases including heatmaps of Plasmodium species correlations, peak malaria incidences by month and year and visualization of year-wise infections, medication status and follow-up rates using Python, Plotly and R Studio.

**Findings:** Among 317,210 suspected malaria patients 77,995 (24.58%) were microscopy-confirmed cases: *Plasmodium vivax (P.v)* (72.52%), *Plasmodium falciparum (P.f)* (24.78%) and mixed infections (2.67%). Out of these, 99.22% received treatment, while 0.77% did not take medication despite tested positive. Follow-up rates on day 7 were low (12.48%), highlighting a significant gap in post-treatment monitor. Hypertension was observed in 39.60% of malaria patients with mixed infections exhibits the highest prevalence 98.32%, followed by *P. falciparum* 75.54% and *P. vivax* 25.16%. Notably, mixed infections are associated with severe clinical outcomes. Parasitaemia levels were highest in *P. falciparum* (1,668,824/µl), followed by mixed infections (78,986/µl) and *P. vivax* (56,694/µl). Over the study period, *P. vivax* incidence rose from 53.39% in 2005 to 86.95% in 2022, while *P. falciparum* infections declined from 45.15% in 2005 to 10.81% in 2016, reflecting advancements in malaria control measures.

**Interpretation:** Deficient treatment completion and insufficient follow-up significantly hinder malaria control in India. Strengthen patient tracking mechanisms and ensuring medication adherence, especially among vulnerable populations are crucial to reduce malaria recurrence and prevent resistance. The decline trend in *P. falciparum* infections underscores progress in control efforts, but the rising incidence of *P. vivax* and mixed infections warrants further investigation and targeted interventions.

## Introduction

Malaria is a life-threatening blood disease caused by *Plasmodium* parasites, which infect the red blood cells (RBCs) and the parasites transmitted to humans through bites of *Plasmodium*-carrying female *Anopheles* mosquitoes. Malaria is preventable and treatable disease if diagnosed and treated in time.^1^ Worldwide, an estimated 247 million cases of malaria occurred in 85 countries, with 619,000 related deaths in 2021.^2^ Malaria is generally more susceptible in infants, children under 5 years, pregnant women, vulnerable travellers, persons with partial immunity, such as cancer patients, and those with immune deficiencies, including HIV/AIDS are at more liability.^3^ The WHO South-East Asia Region (SEARO) ranks second globally in malaria burden, reported 2,614,500 cases (10.5%) and 747,000 deaths (3%), trailing behind the WHO African Region, which accounts for 93.6% (234 million) of global malaria cases and 95.4% (570,000) of malaria-related deaths.^3^ Nine endemic countries in the WHO South-East Asia Region are Bangladesh, Bhutan, DPR Korea, India, Indonesia, Myanmar, Nepal, Thailand, and Timor-Leste account for an estimated 5.4 million cases and 108,000 (2%) malaria-related deaths globally.^2^

India alone contributes 4.26 million cases (79%) of malaria in the South-East Asia Region, with *P. vivax* single attribute for 40% of malaria burden.^2^ In past two decades, South-East Asia region has brought down malaria cases from 17.4 million in 2000 to 4.12 million in 2021, a 76.3% reduction. Additionally, the incidence rate per 1,000 population at risk has deprecated by 82.1%.^2^ Despite these achievements, urban malaria transmission persists due to abandoned and intermittently used wells, partially filled rainwater storage habitats during rainy seasons, water accumulation associated with construction activities and other anthropogenic factors have facilitated the proliferation of major urban malaria vector *Anopheles stephensi*,^4^ thereby contributing to local malaria transmission.^5^ Evaluate existing malaria control strategies for their efficacy and limitations in eliminating transmission, particularly in the context of urban malaria epidemiology. India has significantly contributed to malaria cases in South-East Asia region and is committed to eliminate the disease by 2030.^6,7^ Need to examine the current malaria burden in India, with a particular emphasis on the role of *P. vivax* and the challenges posed by asymptomatic and low-density infections. Elimination may not be possible with the currently available tools and to achieve the ambitious goals by 2030, there is an urgent need for breakthroughs in compulsory treatment for all infected patients and mandatory follow-up on day 7 to block parasite infection and prevent subsequent transmission.

This study aims to analyse the clinical presentation and epidemiological trends of malaria in Mangalore over two decades (2002–2022), highlighting treatment adherence challenges, the rising prevalence of *Plasmodium vivax* cases, the declining of *Plasmodium falciparum* cases and the implications for malaria elimination efforts in India.

### Study site, design and population

Mangalore, officially known as Mangaluru is a coastal city in the southwestern region of Karnataka, India. Geographically nestled between the Laccadive Sea and the Western Ghats, Mangalore enjoys a distinctive ecological setting. It lies approximately 14 km south of the Karnataka-Kerala border, 297 km from Goa and about 352 km west of Bengaluru, the state capital. The city has been designated as one of India’s 100 Smart Cities and is only city in Karnataka that has all four major transportation modes: air, road, rail, and sea.^8^ Situated at an average elevation of 45 meters above sea level, having tropical climate with an annual average temperature of 27°C, humidity levels exceeding 78% (maximum 93% in July) and an average annual rainfall of 3,783 mm, 95% of which occurs during the monsoon season.^9^ These environmental factors, combined with urban settlements near stagnant water bodies, agricultural irrigation practices and suboptimal housing and limited healthcare access, create ideal conditions for high vector densities, significantly contribute to malaria transmission.^5^

A retrospective hospital-based clinical study was conducted over a 21-year period (January 2002 to December 2022) at Wenlock District Government Hospital in Mangalore. This government tertiary care centre is the city’s largest public health facility and serves as the principal referral hospital for malaria diagnosis and treatment in the surrounding region.^10^ It offers free medical care and meals to economically disadvantaged individuals^11^ and their attendants.^5^ The hospital, affiliated with Kasturba Medical College (KMC), Mangalore, has a capacity of approximately 1,000 beds and serves patients from neighbouring districts.^12^ As a hub for medical tourism, Mangalore also receives patients from abroad.^13^ Inclusion criteria for this study were individuals presenting with axillary temperatures >37.5°C and confirmed malaria diagnosis through expert microscopy. Rapid diagnostic tests (RDTs), were included for diagnosis and conformation of malaria,^14^ commercially available RDTs from various manufacturers were utilized from 2013 onward. ^15^

In total, 77,995 patients aged from 1 to 80 years with confirmed malaria infections were enrolled in the study. Among them, 11,700 were adolescents (≤18 years; 15.0%), 46,797 were adult males (>18 years; 60.0%), 19,208 were adult females (>18 years; 24.62%), and 290 identified as transgender individuals (>18 years; 0.37%). Pregnant women, infants (<1 year) and individuals above 80 years were excluded.

The study participants represented a wide demographic including regional ethnic groups of Tuluva, Kannada, and Byari Muslims from Mangalore, Udupi, Hassan, North Kannada, Hubli, and Dharwad^10^. Additionally, the study cohort included Indo-Aryan and tribal populations such as Bagata, Rajuar, Bodo, and Kharia from various Indian states (e.g., Andhra Pradesh, Tamil Nadu, Kerala, Odisha, West Bengal, Jharkhand, Assam, Tripura, Maharashtra, Uttar Pradesh, Bihar). Many of these individuals were engaged in construction, hospitality, and other labour-intensive sectors. Furthermore, participants from Nepal, primarily engaged as private security personnel and domestic service in Mangalore, were also included. Ethnic and regional identification was based on language, food practices, residential history and socio-cultural background.

According to the 2021 Indian Census, Mangalore had a population of 724,159, ^16^ with 1.55% lived in slum areas. In addition to the native population, 10% comprised students pursuing higher education in fields such as science, medicine, commerce and engineering, accompanied by family members and visitors. Moreover, 1.5% religious pilgrims, 5% business-sector workers and 12% worked in different sectors such as construction, domestic service and transportation.

### Ethical statement

The institutional review board of ICMR-National Institute of Malaria Research, New Delhi, India reviewed and approved the study (Ref. No-ECR/NIMR/EC/2012/39). Additionally, ethical clearance was granted by the Research and Ethics Committee of Kasturba Medical College (KMC), Manipal Academy of Higher Education, Udupi, Karnataka, India (Ref. No-IEC KMC MLR 03 16/49). All essential regulatory protocols and ethical guidelines were strictly followed.

### Malaria diagnosis and quantification of parasites Microscopy

Between 2002 to 2022 a total of 3,17,210 clinical cases were screened at Wenlock District Government Hospital Mangalore, India of whom 77, 995 (24.58%) were conformed malaria cases. Among them *Plasmodium vivax (P.v)* 56,578 cases (72.54%), *Plasmodium falciparum (P.f)* 19,332 cases (24.78%) and mixed infections (*P.f + P.v*) 2,085 cases (2.67%).

### Laboratory procedure

Suspected malaria patients were clinically evaluated and confirmed through microscopic examination of peripheral blood smears. Both thick and thin smears were prepared via finger-prick and stained with 4% Giemsa. Slides were air-dried and examined under a Zeiss Primo Star light microscope (Zeiss, Germany) at 1000× magnification (100× oil immersion objective with 10× eyepiece)^10^. Parasite density (parasitaemia) was quantified using the standard methods^11^, ensuring accurate enumeration of parasite density in peripheral blood samples. Clinical parameters were reassessed, and follow-up smears were prepared on day 7 post-treatment to assess drug response.

### Parasitaemia Evaluation

Parasitaemia was assessed in peripheral blood smears of clinically diagnosed patients by three expert microscopists on the first day of hospital admission. Parasitaemia result is mentioned on Table-1.

### Antimalarial treatment and follow-up

#### Therapeutic Significance and Historical Context

Antimalarial medications are crucial antiparasitic agents both synthetically produced and naturally derived, utilized to treat and prevent malaria. These medications primarily target young children and pregnant women, who are the most susceptible to severe malaria in endemic regions. According to the World Health Organization,^17^ young children face a high risk of severe malaria. In contrast, repeated malaria infections in older children and adults can lead to partial immunity, reducing their risk of severe disease.^18^ Pregnant women remain particularly vulnerable to malaria with significant risks to both maternal health and fetal development.^19^

Most of antimalarial drugs are designed to act against the erythrocytic or blood stage of malaria infection which is responsible for the symptomatic phase of the disease. The pre-erythrocytic (hepatic) stage of *Plasmodium* infection remains comparatively less understood in terms of drug action.^20^ Among the prominent antimalarial drugs in current use, two plant-derived compounds have played pivotal roles in malaria management. Quinine, extracted from the bark of the *Cinchona* tree native to South America, was first documented as a remedy in the 16th and 17th centuries. Artemisinin, isolated from *Artemisia annua L.* (Qinghao plant) has been used in traditional Chinese medicine since the 4th century.^21^

These plant-derived compounds have been pivotal in the fight against malaria. Quinine, historically significant, was one of the first treatments for malaria, while Artemisinin and its derivatives have become the cornerstone of modern antimalarial therapy due to their rapid action and efficacy. Despite these advancements in treatment, the challenge of drug resistance remains necessitating sustained research efforts and development of novel antimalarial agents to ensure effective management and long-term malaria control.

#### Treatment guide line for clinically conformed positive malaria

Prompt and accurate diagnosis facilitated the administration of targeted antimalarial therapies according to *Plasmodium* species identified in clinically confirmed malaria cases. For patients infected with uncomplicated *Plasmodium vivax* the regimen comprised oral Chloroquine administered over 3 days, followed by a 14-day course of Primaquine to achieve radical cure and prevent relapse. Infections due to *Plasmodium falciparum* were managed with a standard artemisinin-based combination therapy (ACT), consisting of Artesunate, Sulfadoxine and Pyrimethamine for 3 days, supplemented by a single dose of Primaquine on the second day to interrupt transmission.

Patients diagnosed with mixed infections (*P. falciparum* + *P. vivax*) received ACT for 3 consecutive days along with Primaquine 0.25 mg/kg body weight daily for 14 days^14^, ensuring coverage against both species and minimizing the risk of recrudescence or relapse.

Close monitoring of drug adherence and follow-up were systematically implemented across all three groups to reinforce treatment efficacy and optimize patient outcomes. These regimens reflect current best practices tailored to species-specific malaria profiles and underscore the importance of stratified therapeutic approaches in endemic settings.

#### Antimalarial Medication Dispensation

Prior to dispense antimalarial medication, laboratory technicians at malaria clinic conducted brief pre-dispensation counselling to clinically diagnosed malaria patients. These sessions emphasized adherence to advised medication, importance of follow-up on day 7 and need to consult with attending physicians to monitor treatment response and detect potential complications. Such patient instructions were intended to enhance recovery outcomes and mitigate disease progression.

Following medication dispensation, patient behaviours varied considerably. While some individuals proceeded to clinical consultation as advised, others initiated their treatment independently at home, relying solely on technician-provided instructions and bypassing medical supervision. In complicated malaria cases, physicians recommended hospital admission for parenteral therapy and close monitoring. However, several admitted patients absconded during treatment. Potentially due to fear of hospitalization, perceived financial burdens, emotional distress and witness critically ill co-patients.

Operational barriers included absence of accompany caregivers, lack of reliable contact information and non-cooperation from patients, even among those who initially consented to follow-up. These challenges impeded continuity of care, completion of antimalarial regimens and increased the risk of treatment failure, potential recrudescence and sustained community transmission.

#### Statistical Analysis

To compare the exposure, outcome and categorical clinical characteristics among the three groups i.e. *Plasmodium falciparum*, *Plasmodium vivax* and mixed malaria infections were analysed using Fisher’s exact test. We calculated the odds ratios (OR), 95% confidence intervals (CI) and p-value for each clinical characteristic using MedCalc online statistical calculator (https://www.medcalc.org/calc/odds_ratio.php). Data visualization was carried out using RStudio, the Plotly library for R and Julius Artificial Intelligence (AI) tools to ensure valid and accurate data representation of variables such as antimalarial medication adherence, follow-up compliance, and treatment initiation. The Pearson correlation coefficients were computed using R and Python-based statistical frameworks. Data visualization was conducted using RStudio (https://rstudio.com/products/rstudio/download/) and Plotly for R (https://plotly.com/r/), enabling generation of correlation heatmaps and matrix plots. Julius AI-enhanced graphical tools were employed to augment the precision and readability of visual outputs, ensuring accurate representation of analytical results.

## Results

### Clinical Malaria Incidence in 2002 to 2022

Clinical malaria cases from 2002 to 2022 were depicted in the line graph, using Julius Artificial intelligence tools, with the temporal tends illustrated in Figure 1. The line graph employs distinct colour codes to represent the following parameters are outer blue line indicates total number of blood samples collected for malaria diagnosis, the violet line shows total malaria-positive cases, the orange line represents number of *P. vivax* cases, the green line shows number of *P. falciparum* cases and the red line indicates number of mixed infections (*P. falciparum* + *P. vivax*). The highest annual malaria prevalence was observed in 2014 with 23,123 cases, followed by 22,219 cases in 2005, 20,481 cases in 2006 and 20,097 cases in 2015. The peak malaria incidence rates were 39.43% (7,443/18,872) recorded in 2004, followed by 36.63% (8,139/22,219) in 2005, 33.78% (6,920/20,481) in 2006 and 29.26% (6,767/23,123) in 2014. The predominant number of *P. vivax* malaria infections was recorded in 2013 with 86.82% (3,994/4,600) of cases, followed by 80.12% (5,422/6,767) in 2014, 76.59% (2,694/3,517) in 2003 and 64.12% (4,773/7,443) in 2004. The maximum number of *P. falciparum* cases was 45.15% (3,675/8,139) cases observed in 2005, followed by 40.92% (2,832/6,920) in 2006, 34.65% (1,443/4,164) in 2007 and 26.01% (1,180/4,536) in 2009. The most significant number of mixed infections peaked 11.55% (511/4,425) in 2015, followed by 6.07% (191/3,144) in 2016, 5.17% (350/6,767) in 2014 and lowest 6.14% (15/244) in 2021. These findings highlight the fluctuations of epidemiological dynamics in malaria over the two-decade period with significant variances in the prevalence and types of infections imparted in Figure 1.

**Figure 1:**
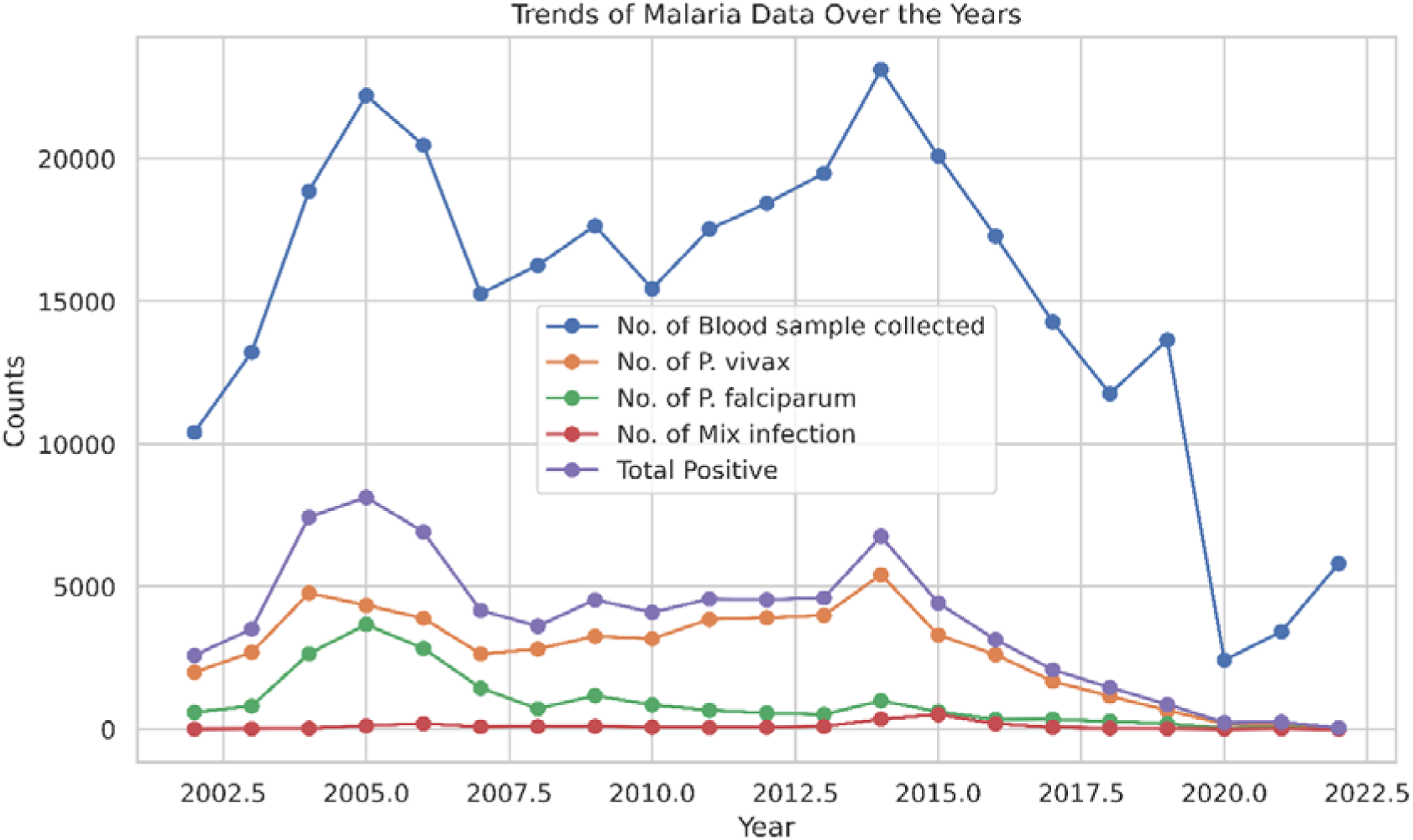
A line graph showing the annual number of clinically reported malaria cases in Mangalore, India from 2002 to 2022. The X-axis represents the years, and the Y-axis represents the number of cases. The graph displays the highest number of infections is *P. vivax*, followed by *P. falciparum*, and then mixed infections (*P. falciparum* + *P. vivax*).

### 21 Years of Clinical Malaria Epidemiology

From 2002 to 2022, in Mangalore, microscopy-based conformed malaria cases were reported 77,995 cases (positive rate; 24.58%) out of 3,17,210 suspected cases. Of these *Plasmodium vivax* (*P.v*) attributed to 56,578 cases (positive rate; 72.54%), *Plasmodium falciparum* (*P.f*) to 19,332 cases (positive rate; 24.78%) and mixed infections cases identified as 2,085 cases (positive rate; 2.67%). The 21-year clinical findings underscore the epidemiological dominance of *Plasmodium vivax* (*P.v*) in the region, the substantial presence of *Plasmodium falciparum* (*P.f*) and the sporadic but significant occurrence of mixed infections, provide critical insights into malaria case management strategies, as illustrated in the statistical pie-chart presentation using Julius AI generated in Figure 2.

**Figure 2:**
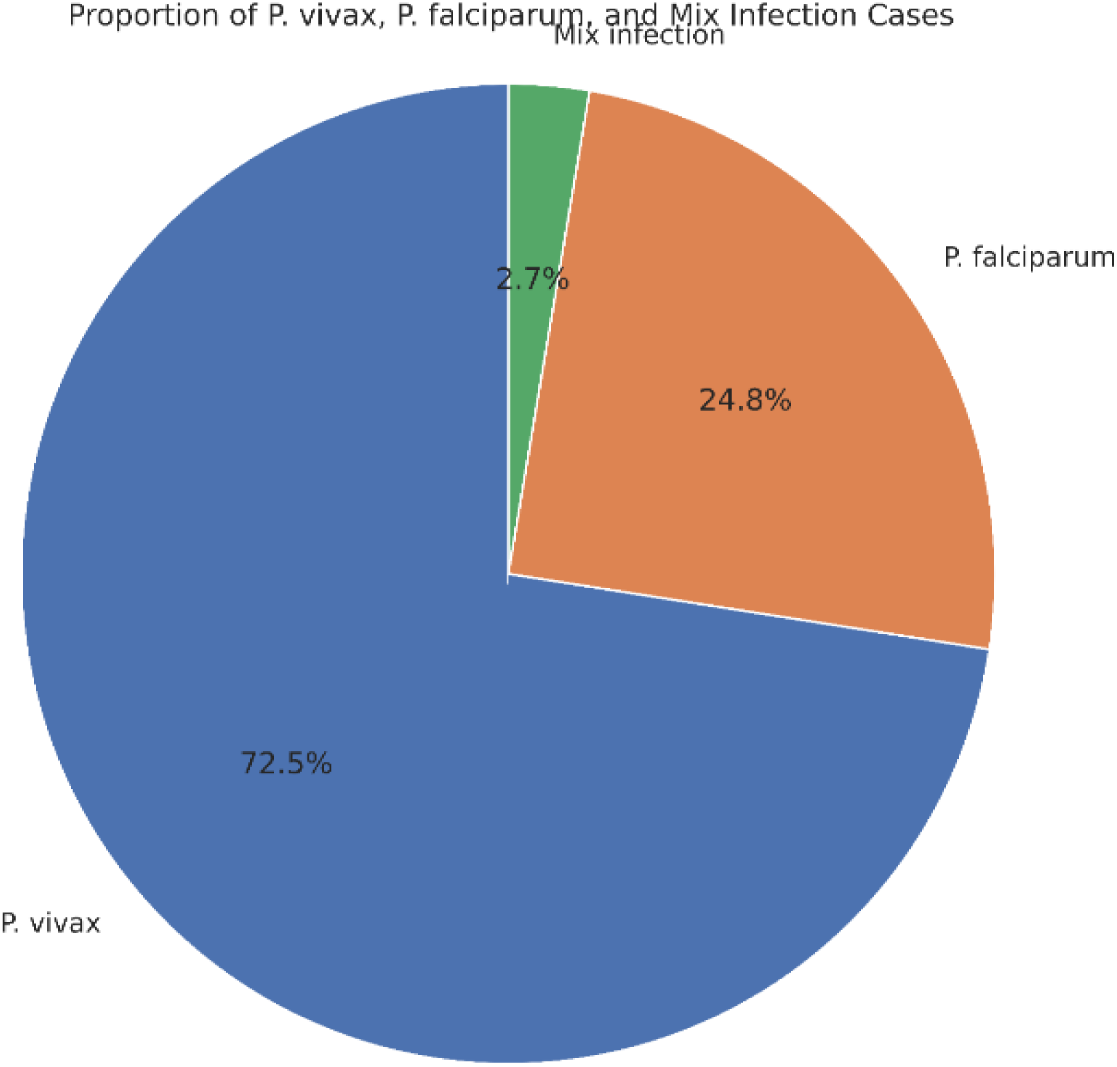
21-years of clinical malaria pi-chart generated applying Julius Artificial Intelligence tools, manifest the result of *P. vivax*, *P. falciparum* and mixed malaria infection cases burden. In Pi-chart, blue colour represents *P. vivax* cases (72.5%), orange colour denominated *P. falciparum* (24.8%) and green colour performed mixed infection (2.7%) cases.

### Clinical Characteristics of Malaria Appearance

In a cohort of 77,995 clinically confirmed malaria cases in Mangalore encompasses with *Plasmodium vivax, Plasmodium falciparum* and mixed form of malaria, a spectrum of clinical manifestations and comorbidities was documented in Table 2. The analysis revealed distinct symptom profiles across species are hypertension was most prevalent in mixed infections 98.32% (2,050/2,085), moderate level in *P. falciparum* 75.54% (14,604/19,332) and least in *P. vivax* cases 25.15% (14,233/56,578), yielding an overall prevalence 39.60% (Odds ratio: 1.0491; CI 95%:1.0164 to 1.0828; *P* value^a^:0.0030). General febrile temperature was near-universal, reported 100% (2,085/2,085) in mixed infections, 99.87% (56,507/56,578) in *P. vivax* cases and 99.83% (19,300/19,332) in *P. falciparum* cases. Nausea was observed 97.09% (18,771/19,332) in *P. falciparum*, 95.92% (2,000/2,085) in mixed infections and 64.96% (36,756/56,578) in *P. vivax* cases. Vomiting was most common in mixed infection 62.15% (1,296/2,085), followed by *P. falciparum* 40.79% (7,887/19,332) and *P. vivax* 16.59% (9,393/56,578). Loss of appetite/anorexia was more prevalent in *P. falciparum* 76.49% (14,788/19,332), in mixed infections 72.37% (1,509/2,085) and in *P. vivax* 38.69% (21,887/56,578). Breathing difficulty was reported in mixed infection 47.43% (989/2,085), *P. falciparum* 40.80% (7,889/19,332) and *P. vivax* 12.01% (6,799/56,578). Burning sensation in the eyes was most frequent in *P. falciparum* cases 7.23% (1,399/19,332), followed by mixed infection cases 4.70% (98/2,085) and lowest in *P. vivax* cases 1.25% (708/56,578). Burning sensation in the chest was most frequent in mixed infection 23.40% (488/2,085), compared to *P. falciparum* 13.43% (2,597/19,332) and lowest) in *P. vivax* 1.78% (1,008/56,578).

**Table 1:** In *P. falciparum* malaria has highest number of parasitaemia 1,668,824/µl and in average number of parasites 834,512/µl but in *P. vivax* has lowest 56,694/µl and in average number of parasites 28,527/µl.

| Table 1 Clinically Diagnosed *P. falciparum*, *P. vivax* and Mixed Infection Parasitaemia Report |  |  |  |
| --- | --- | --- | --- |
| Type of Infection | Parasite Range (Parasites/μl) | Average (Parasites/μl) | Remarks |
| *P. falciparum* | 200 – 1,668,824 | ~ 834, 512 | Highest Parasitaemia recorded |
| Mixed (*P.f* + *P.v*) | 520 – 78,986 | ~ 39,753 | Second Highest levels |
| *P. vivax* | 360 – 56,694 | ~ 28,527 | Lowest observed levels |

**Table 2:** Clinical malaria data represented as number of patients (percentages calculate i.e. *total number of infected patients and all other percentages were based on number of patients in each infected group) A p-value of less than 0.05 was considered statistically significant in this study, suggesting that the differences observed between the groups were unlikely to be due to random variation alone.

| Table 2 Clinical Characteristics of *P. falciparum*, *P. vivax* and Mixed malaria Infection |  |  |  |  |  |  |  |
| --- | --- | --- | --- | --- | --- | --- | --- |
| Characteristics | *P. vivax*<br>(n =<br>56,578) | *P. falciparum*<br>(n =<br>19,332) | Mixed Infection<br>(n =<br>2,085) | Overall<br>(n =<br>77,995) | Odds ratio | CI 95% | *P* value<sup>a</sup> |
| **Fever** | 56,507<br>(99.87%) | 19,300<br>(99.83%) | 2,085<br>(100%) | 77,892<br>(99.86%) | 0.1247 | 0.1219 to<br>0.1275 | 0.0001 |
| **Headache** | 56,500<br>(99.86%) | 19,330<br>(99.98%) | 2,081<br>(99.80%) | 77,911<br>(99.89%) | 0.1251 | 0.1223 to<br>0.1279 | 0.0001 |
| **Chills** | 50,100<br>(88.55%) | 18,898<br>(97.75%) | 2,080<br>(99.76%) | 71,078<br>(91.13%) | 0.1516 | 0.1482 to<br>0.1552 | 0.0001 |
| **Shivering** | 17909<br>(31.65%) | 14856<br>(76.84%) | 2017<br>(96.73%) | 34,782<br>(44.59%) | 0.7024 | 0.6817 to<br>0.7237 | 0.0001 |
| **Sweating** | 53,878<br>(95.22%) | 18,000<br>(93.10%) | 1900<br>(91.12%) | 73778<br>(94.59%) | 0.1197 | 0.1170 to<br>0.1226 | 0.0001 |
| **Cough** | 11,177<br>(19.75%) | 10,167<br>(52.59%) | 788<br>(37.79%) | 22132<br>(28.37%) | 0.8329 | 0.8023 to<br>0.8645 | 0.0001 |
| **Running nose** | 4002<br>(7.07%) | 3,980<br>(20.58%) | 489<br>(23.45%) | 8471<br>(10.86%) | 0.9914 | 0.9333 to<br>1.0530 | 0.7783 |
| **Nausea** | 36,756<br>(64.96%) | 18,771<br>(97.09%) | 2,000<br>(95.92%) | 57527<br>(73.75%) | 0.2737 | 0.2671 to<br>0.2805 | 0.0001 |
| **Vomiting** | 9,389<br>(16.59%) | 7,887<br>(40.79%) | 1,296<br>(62.15%) | 18,572<br>(23.81%) | 0.7219 | 0.6930 to<br>0.7521 | 0.0001 |
| **Sore throat** | 7,998<br>(14.13%) | 3191<br>(16.50%) | 188<br>(9.01%) | 11,377<br>(14.58%) | 0.1647 | 0.1555 to<br>0.1744 | 0.0001 |
| **Bitter mouth** | 14,988<br>(26.49%) | 6,090<br>(31.50%) | 500<br>(23.98%) | 21,578<br>(27.66%) | 0.1729 | 0.1659 to<br>0.1802 | 0.0001 |
| **Fatigue** | 24,880<br>(43.97%) | 11,865<br>(61.37%) | 1277<br>(61.24%) | 38022<br>(48.74%) | 0.2396 | 0.2325 to<br>0.2470 | 0.0001 |
| **Dizziness** | 23,455<br>(41.45%) | 8,876<br>(45.91%) | 899<br>(43.11%) | 33,230<br>(42.60%) | 0.1519 | 0.1468 to<br>0.1571 | 0.0001 |
| **Abdominal pain** | 12,077<br>(21.34%) | 5988<br>(30.97%) | 576<br>(27.62%) | 18,641<br>(23.90%) | 0.2572 | 0.2464 to<br>0.2685 | 0.0001 |
| **Diarrhoea** | 2086<br>(3.68%) | 183<br>(0.94%) | 23<br>(1.10%) | 2292<br>(2.93%) | 0.0086 | 0.0070 to<br>0.0106 | 0.0001 |
| **Loss of appetite/anorexia** | 21,887<br>(38.68%) | 14,788<br>(76.49%) | 1509<br>(72.37%) | 38,184<br>(48.95%) | 0.4701 | 0.4567 to<br>0.4839 | 0.0001 |
| **Myalgia (back and limbs)** | 7,893<br>(13.95%) | 8371<br>(43.30%) | 986<br>(47.29%) | 17250<br>(22.11%) | 1.1177 | 1.0714 to<br>1.1659 | 0.0001 |
| **Breathing difficulty** | 6799<br>(12.01%) | 7889<br>(40.80%) | 989<br>(47.43%) | 15677<br>(20.10%) | 1.3227 | 1.2652 to<br>1.3829 | 0.0001 |
| **Burning sensation in eyes** | 708<br>(1.25%) | 1399<br>(7.23%) | 98<br>(4.70%) | 2205<br>(2.82%) | 3.6792 | 3.2483 to<br>4.1672 | 0.0001 |
| **Burning sensation in chest** | 1008<br>(1.78%) | 2597<br>(13.43%) | 488<br>(23.40%) | 4093<br>(5.24%) | 5.3129 | 4.8295 to<br>5.8448 | 0.0001 |
| **Hypertension** | 14,233<br>(25.15%) | 14,604<br>(75.54%) | 2050<br>(98.32%) | 30887<br>(39.60%) | 1.0491 | 1.0164 to<br>1.0828 | 0.0030 |

### Distribution and Epidemiological Trend of *Plasmodium vivax* malaria

The annual proportion of *Plasmodium vivax* cases among total confirmed malaria infections in Mangalore from 2002 to 2022 is depicted in Figure 3. The analysed data reveal a marked upward trend in *P. vivax* prevalence, despite an overall decline in total malaria incidence. The highest prevalent of *P. vivax* malaria was recorded in 2022 86.95% (40/46), followed by an incidence rate 86.82% (3994/4600) in 2013. In 2012, infection rate was 86.14% (3911/4540), which decreased to 84.43% (3855/4566) in 2011 and further to 83.11% (2613/3144) in 2016. By 2020, the prevalence rate declined to 81.05% (184/227), with a rate of 80.49% (1676/2082) in 2017, 80.12% (5422/6767) in 2014, 79.27% (1159/1462) in 2018, 77.62% (2810/3620) in 2008, 77.49% (3175/4097) in 2010, 77.07% (1994/2587) in 2002, 76.75% (667/869) in 2019, 76.59% (2694/3517) in 2003, 74.69% (3305/4425) in 2015, 71.84% (3259/4536) in 2009, 68.03% (166/244) in 2021, 64.12% (4773/7443) in 2004, 63.40% (2640/4164) in 2007, 56.28% (3895/6920) in 2006, and 53.39% (4346/8139) in 2005. The incidence of *P. vivax* infections increased significantly from 53.39% in 2005 (4,346/8,139) to a peak of 86.95% in 2022 (40/46), indicating rise in the proportion of *P. vivax* cases relative to the overall decline in total malaria incidence. This trend highlights the growing threat of *P. vivax*, which has implications for policy, case management, economic impact and the effectiveness of malaria control measures over the observed period from 2002 to 2022.

**Figure 3:**
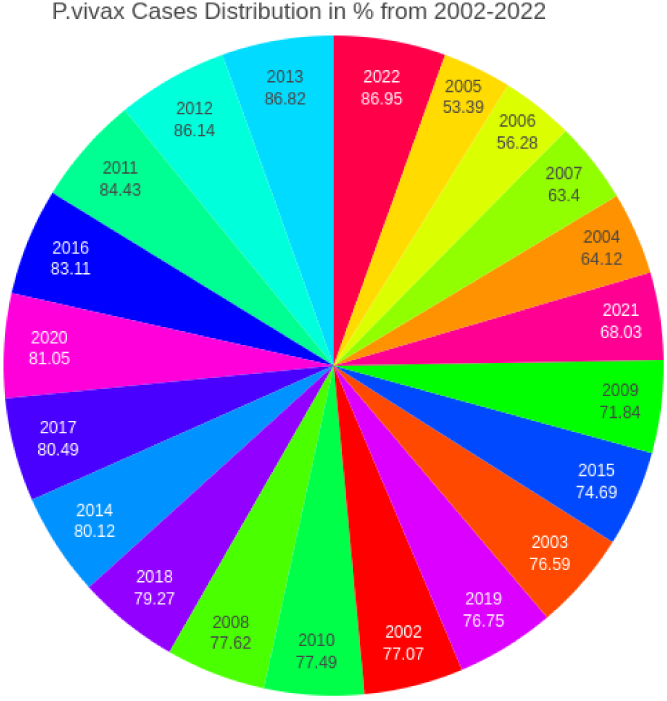
In pi-chart represents the percentage of *P. vivax* cases from 2002 to 2022. The highest vivax cases show in the year 2022, red in colour and lowest in the year 2005, yellow in colour.

### Distribution of *Plasmodium falciparum* malaria

The highest number of *Plasmodium falciparum* infections was recorded in 2005, with a prevalence rate of 45.15% (3,675/8,139), followed by 40.92% (2,832/6,920) in 2006 and 35.53% (2,645/7,443) in 2004. A progressive decline was observed thereafter, with prevalence rates dropping to 34.65% (1,443/4,164) in 2007 and further to 26.01% (1,180/4,536) in 2009. By 2021, the prevalence cases declined to 25.81% (63/244), with a rate of 23.03% (810/3517) in 2003, 22.69% (587/2587) in 2002, 21.4% (186/869) in 2019, 20.82% (853/4097) in 2010, 19.80% (717/3620) in 2008, 18.67% (273/1462) in 2018, 16.62% (346/2082) in 2017, 14.41% (658/4566) in 2011, 13.76% (609/4425) in 2015, 12.42% (564/4540) in 2012, 11.13% (512/4600) in 2013, 10.81% (340/3144) in 2016. In 2022 *P.f* accounted 13.04% (6/46) with notably low rates in precedes years such as 16.74% (38/227) in 2020, 14.70% (995/6767) in 2014. The prevalence of *Plasmodium falciparum* infections exhibited a significant decline from a peak of 45.15% (3,675/8,139) in 2005 to 10.81% (340/3,144) in 2016 reflects substantial progress in malaria control measures over the two-decade period from 2002 to 2022. The overall, decline in *P. falciparum* distribution, as illustrated in Figure 4, underscores the impact of sustained vector control interventions, improved diagnostic coverage and targeted treatment strategies.

**Figure 4:**
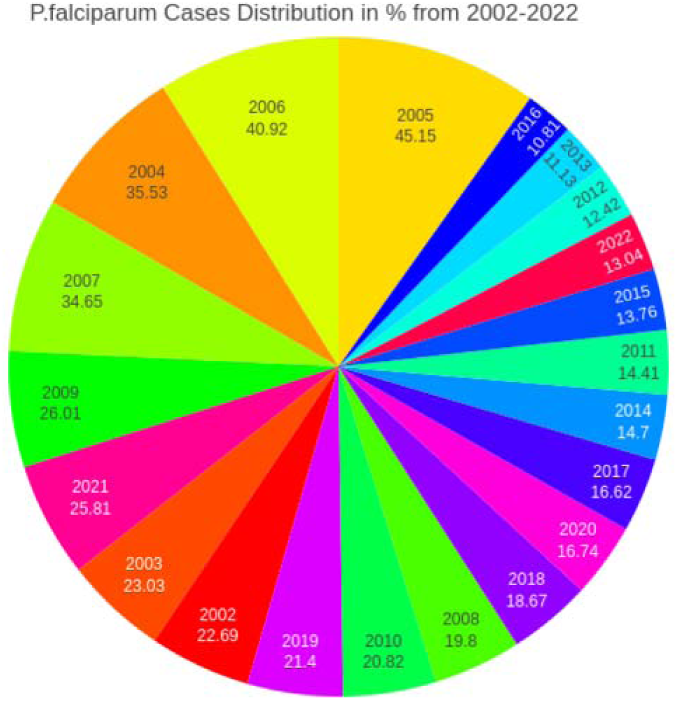
In pi-chart, the different colour coated line represents the percentage of *P. falciparum* cases from 2002 to 2022. The highest *P. falciparum* infection show in the year 2005, represents yellow in colour and lowest in the year 2016, represent blue in colour and no. of cases declined in anticlockwise form.

### Distribution and Clinical Implications of Mixed Infections Malaria

Over a 21-year of clinical study in Mangalore, the incidence of mixed infections exhibited a gradual increase, whereas declining trend of *P. falciparum* mono-infections. The highest prevalence of mixed infections were observed 11.55% (511/4,424) in 2015, 6.14% in 2021, 6.07% in 2016, 5.17% in 2014, 2.88% in 2017, 2.78% in 2006, 2.53% in 2008, 2.2% in 2020, 2.14% in 2009, 2.05% in 2018, 2.04% in 2013, 1.94% in 2007, 1.84% in 2019, 1.68% in 2010, 1.44% in 2005, 1.43% in 2012, 1.16% in 2011, 0.36% in 2003, 0.33% in 2004, 0.23% in 2002, and no infection (0%) in 2022. Mixed infections demonstrated intermittent spikes most notably in 2015 (11.55%), posing diagnostic challenges and complicating case management due to increased clinical severity and resistance risk is delineated in Figure 5. The rising trend in mixed infections poses significant challenges to accurate diagnosis, necessitating the involvement of trained expert microscopists for quality diagnostic accuracy or the implementation of freely available molecular diagnostic/Polymerase Chain Reaction (PCR) setups. Misdiagnosis of mixed infections as either *Plasmodium falciparum* (*Pf)* or *Plasmodium vivax* (*Pv*) can lead to inappropriate treatment, increasing the probability of antimalarial drug resistance by up to 100%, presenting a general threat to effective case management. Therefore, it is imperative to improve current surveillance tools and techniques for malaria elimination. The clinical complications associated with mixed infections are severe, debilitating and chronic compared to those caused by *Plasmodium falciparum* and *Plasmodium vivax* mono-infection. Mixed infections often result in higher rates of severe clinical complications, such as hypertensive comorbidity, gastrointestinal manifestations and respiratory distress. Among all malaria patients, 39.60% had hypertension, with mixed infection patients having the highest rate at 98.32%, followed by *P. falciparum* patients at 75.54%, and *P. vivax* patients at 25.16% [Table 2].

**Figure 5:**
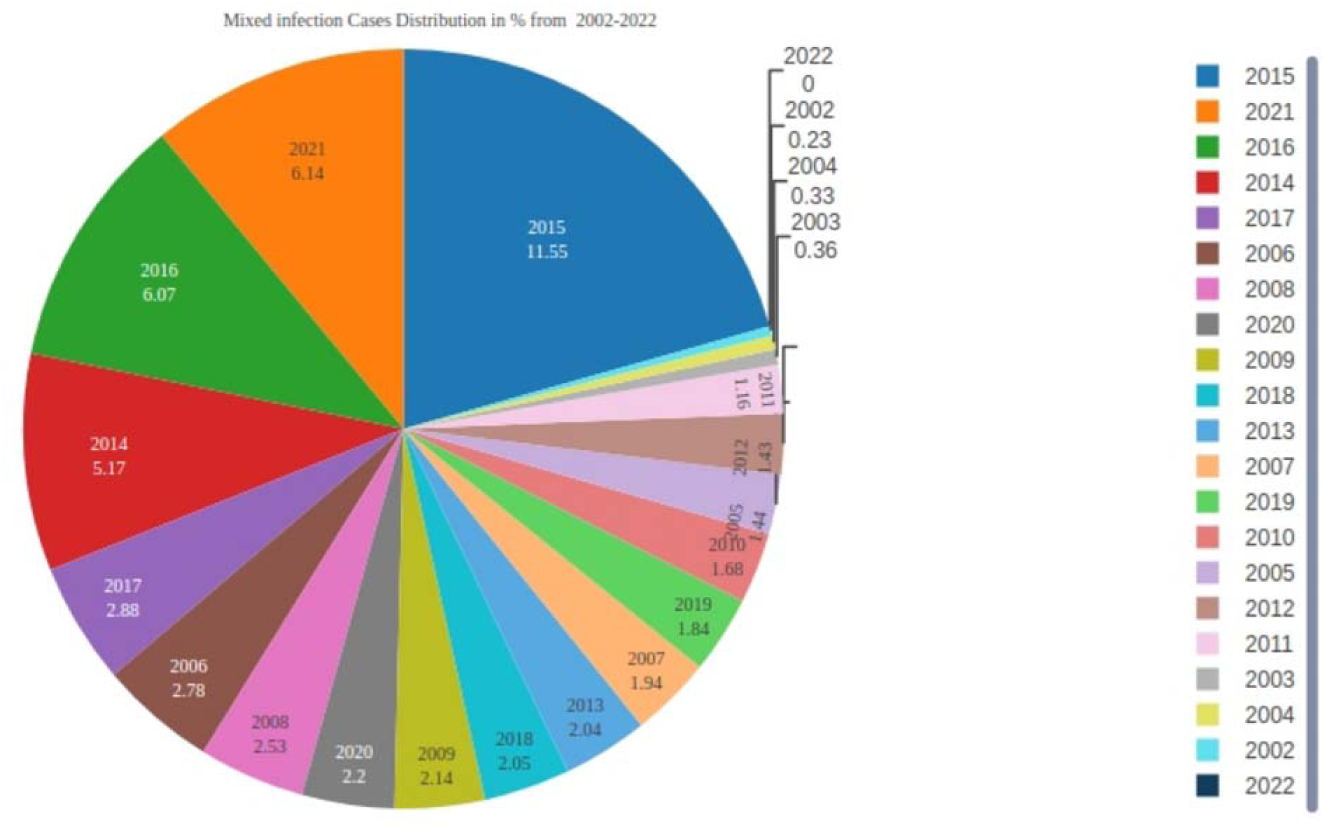
Pi-chart represents the mixed infection cases percentage in 2002 to 2022. The values of infection rate show from highest in 2015, blue in colour to low in 2022, red in colour and decrease the number of infections express in anticlockwise form.

### Distribution of Peak Malaria Cases by Month and Year in 2002 to 2022

In the heatmap, visualize the temporal distribution of peak malaria cases across different months over a 21-year period, from 2002 to 2022. Here is a detailed breakdown of the elements and insights from the heatmap, explained in Figure 6.

**Figure 6:**
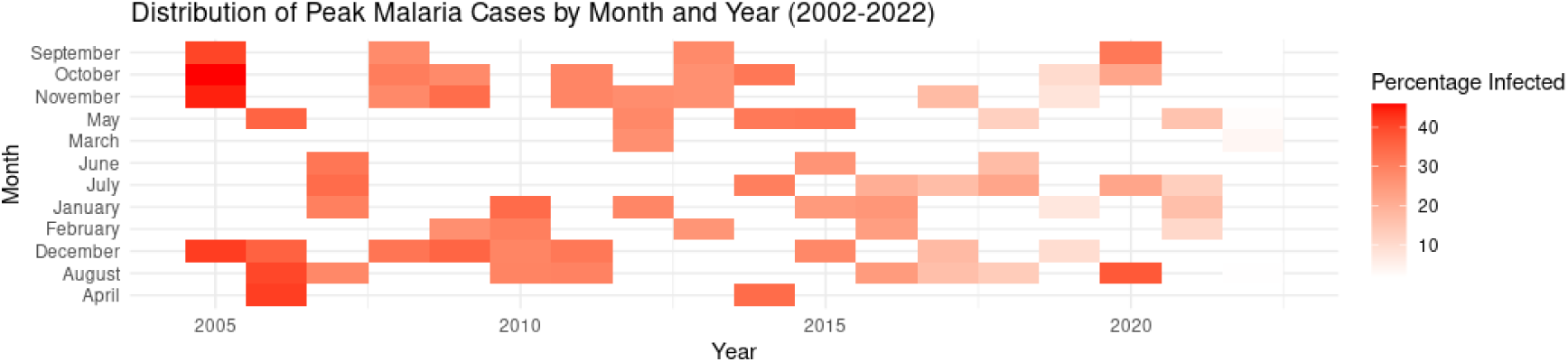
The heatmap distribute the peak malaria cases in month and year from 2002 to 2022. The X-axis represents the year from 2002 to 2022 along with each tick mark corresponds to specific year within this range. Y-axis represents each month of the year from January to December. The months are ordered from September at the top to April at the bottom, indicating the calendar cycle. The percentage of infection rate mentioned on colour scale at the right side of the heatmap.

The heatmap uses a gradient of red shades to represent the percentage of population infected with malaria. White shades indicate the lower percentage of infections, while light red to dark red indicates progressively higher percentages of infections. The darkest red represents the highest peak percentage of infections as shown in the legend on the right.

The heatmap reveals the months and years when malaria cases peaked. Darker red blocks indicate months with higher infection rates in specific years. Observing the seasonal concentration of darker shades helps identify any seasonal trends or patterns in malaria outbreaks. The interannual variability in the distribution and intensity of peaks varies significantly across different years, indicating fluctuations in malaria transmission rates.

Clinical Observation of malaria in 21 years are the high Infection periods in 2004, peak infections occur in December and April, but in 2005, the peak is in September. From 2008 to 2010, there are consistently high infection rates across different months, particularly May and October. From 2015 to 2016, there are prominent peaks in November, with multiple months showing high infection rates. In 2020 to 2021, significant peaks infections occur in July and December. During high infection periods, there is no fixed month for peak malaria infections, it varies from month to month in each year. The low transmission years such as 2012, 2014 and 2018, show fewer or less intense peaks in infections, suggesting potentially lower overall transmission rates during these periods. The seasonal trends of malaria transmission trend to peak from late summer to early winter (September to December) in multiple years, indicating a possible seasonal influence on malaria transmission. The Implications for Research and Public Health are seasonal interventions for public health initiatives could be timed to precede the peak infection months to maximize impact. Resource allocation understands the temporal distribution helps in planning and allocating resources such as distribution of insecticide-treated bed nets, indoor residual spraying campaigns, ensuring availability of diagnostics, antimalarial drugs and vector control tools during high-risk periods. For further research, the heatmap suggests need for further investigation into the causes of temporal variations such as environmental factors (rainfall and temperature), human behaviour and mobility, case management efficacy and vector ecology and breeding dynamics.

In Conclusion, the heatmap provides a visual summary of malaria transmission dynamics over two decades, highlighting both seasonal patterns and year-to-year variability. This valuable information insights for strategic planning, surveillance enhancement and targeted interventions in malaria endemic regions for effective malaria control and prevention measures.

### Correlation Analysis of Malaria Variables by Heatmap Visualization

A heatmap generated using Julius Artificial Intelligence (AI), visualizes Pearson correlation coefficients among key variables in malaria surveillance over a 21-year period depicted in Figure 7. Variables include total blood samples collected, *Plasmodium vivax* cases, *P. falciparum* cases, mixed infections and total malaria-positive cases.

**Figure 7:**
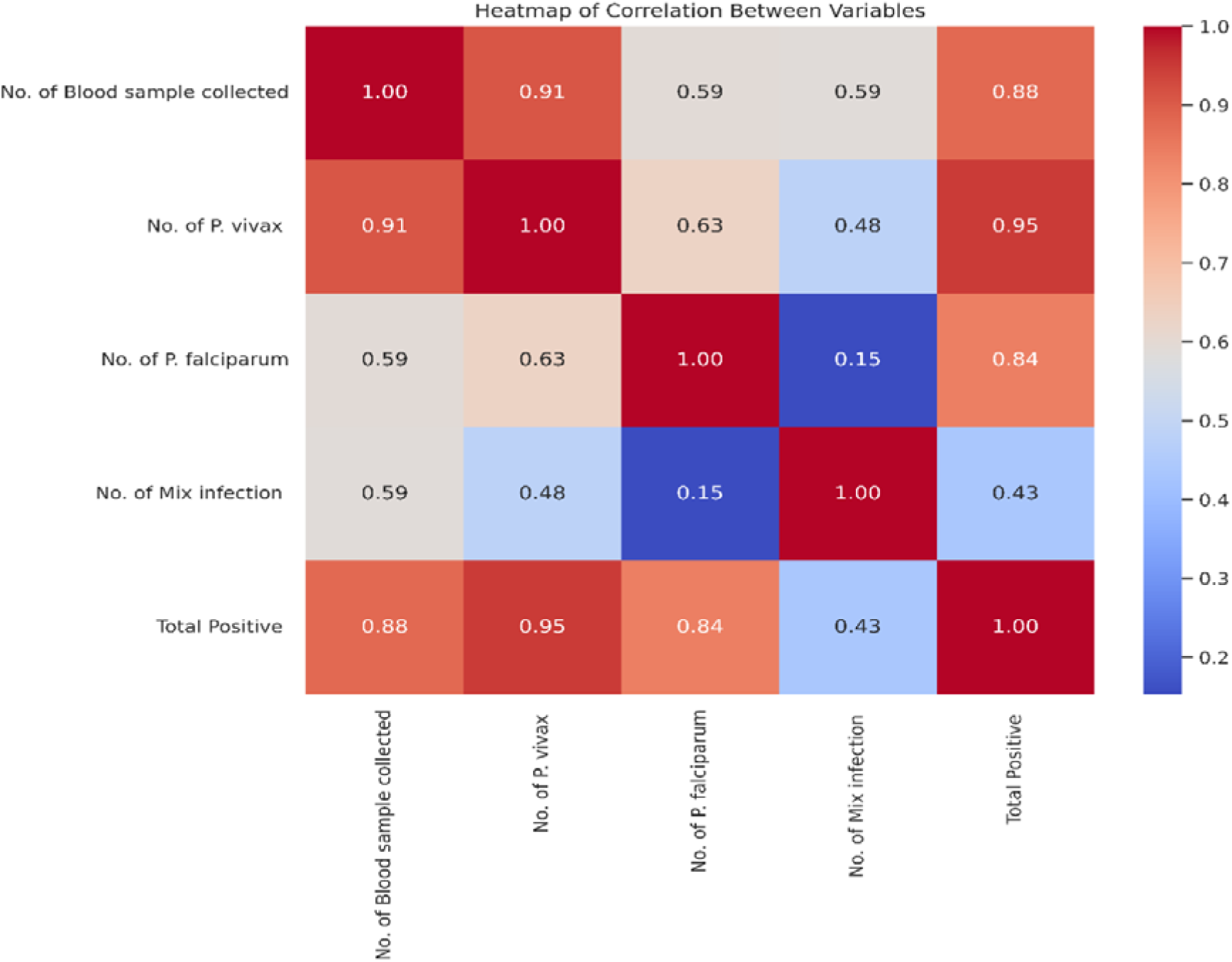
Heatmap generated by Julius Artificial intelligence (AI), represents the correlation between variables in *Plasmodium vivax*, *Plasmodium falciparum* and Mixed infection of malaria from 2002 to 2022. The strong positive correlation demonstrates in *P. vivax* malaria case and low in Mixed infection cases in 21 years clinical research.

Each cell in the heatmap represents a correlation coefficient ranging from –1 to +1, where values near +1 indicate a strong positive correlation (both variables increase proportionally), values near 0 suggest no linear relationship and values near –1 reflect a strong negative correlation (one variable increases as the other decreases). The heatmap’s colour gradient, from dark blue (–1) to dark red (+1) visually encodes the strength and direction of these associations.

Strong positive correlations were observed between *P. vivax* cases and total positive cases (r = 0.95). Blood samples collected and *P. vivax* cases (r = 0.91). Blood samples and total positive cases (r = 0.88). This suggests that *P. vivax* trends are tightly linked with overall malaria surveillance metrics, possibly due to its dominance in the region and seasonal predictability. Moderate correlations included *P. falciparum* cases and total positives (r = 0.84). Blood samples with *P. falciparum* cases and mixed infections (r = 0.59 each). *P. falciparum* shows intermediate correlation, indicating a more variable transmission pattern, possibly influenced by vector ecology, drug resistance or localized outbreaks. Weak correlations were found between Mixed infections and total positive cases (r = 0.43). *P. falciparum* and mixed infections (r ≈ 0). This may reflect their sporadic occurrence, diagnostic challenges or underreporting due to low parasitaemia.

The colour gradient ranges from dark blue (strong negative correlation) to dark red (strong positive correlation), with light blue indicating negligible correlation. These findings suggest that increased blood sampling is strongly associated with higher detection of *P. vivax* and overall malaria positivity, while mixed infections exert minimal influence on total case numbers. This correlation matrix provides actionable insights for optimizing diagnostic strategies and resource allocation in malaria surveillance programs. These patterns underscore species-specific transmission dynamics and inform targeted diagnostic and surveillance strategies.

### Analyses of Patient’s Medication Status

The stacked bar chart in Figure 8 illustrates longitudinal trends in the number of malaria-positive cases that did not receive antimalarial medication, stratified by infection type over a 21-year period (2002–2022). Each bar represents the cumulative annual count of untreated cases with color-coded segments denote specific infection categories are *Plasmodium vivax* (*Pv*; red), *Plasmodium falciparum* (*Pf*; green), mixed infections (*Pv* + *Pf*; blue) and cumulative burden (purple) of total untreated cases. The total height of each bar corresponds to the aggregate number of untreated cases per year, while the purple overlay line traces the overall annual total.

**Figure 8:**
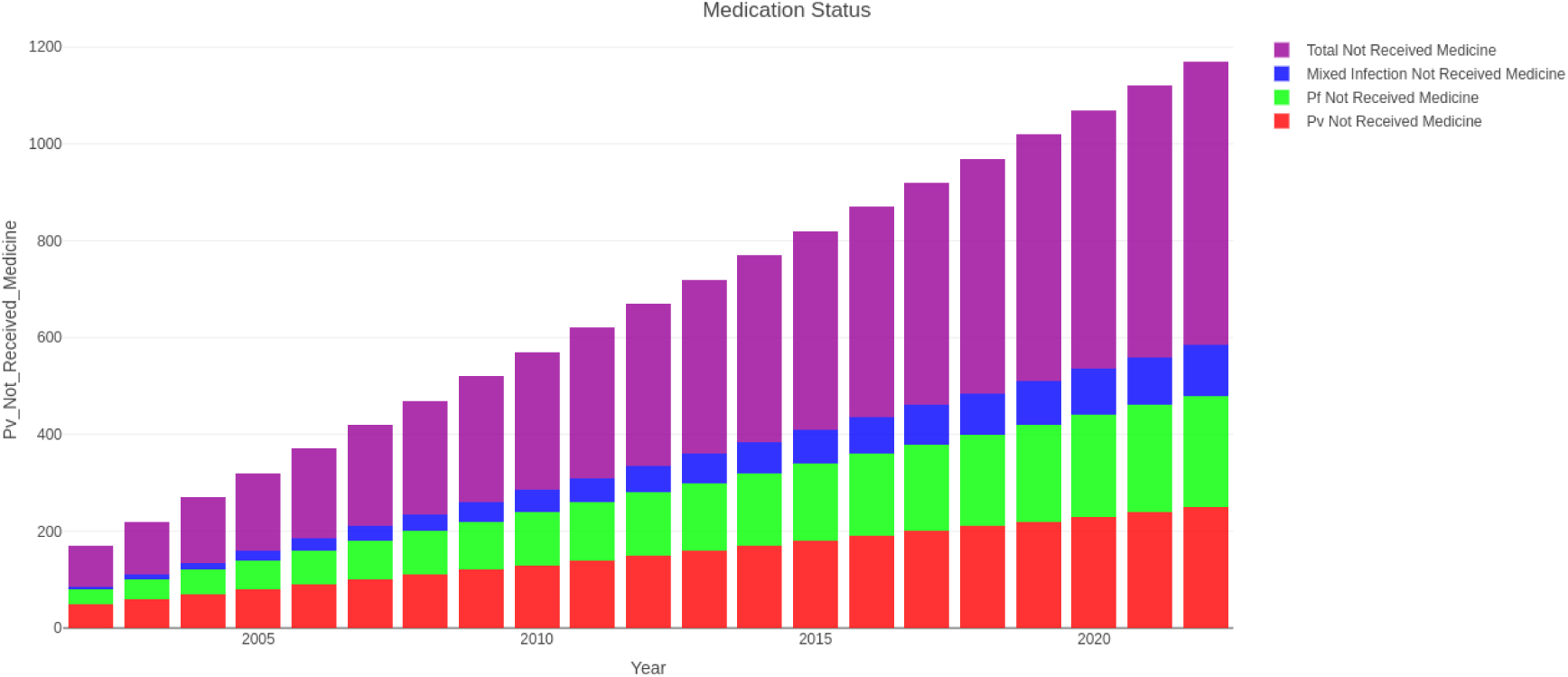
In stacked bar chart show the antimalarial medication status for *P.v*, *P.f* and Mixed infection cases, in stacked bar chart, X-axis shows on the years from 2002 to 2022. Y-axis represents on the number of cases. The stacked bar colour medication status is represented on the right side of the chart.

A progressive increase in the total number of untreated malaria cases is evident across the study period, suggesting a systemic challenge in treatment delivery. The number of untreated *P. vivax* cases exhibits a consistent upward trajectory, represents the largest proportion of untreated cases annually. This trend may reflect diagnostic limitations in detecting low-density *Pv* infections or gaps in follow-up care. Untreated *P. falciparum* cases also show a steady rise, albeit at a lower magnitude compared to *Pv*. Given *Pf* cases higher pathogenicity, this trend raises concerns about potential morbidity and mortality. Mixed infections remain relatively stable but show a modest increase in recent years. Their contribution to the total untreated burden is comparatively lower. The total untreated cases (purple line) demonstrate a marked escalation, underscoring the compounded effect of rising untreated cases across all categories.

The increasing burden of untreated malaria cases poses significant public health risks, including prolonged parasitaemia, increased transmission potential and heightened risk of severe disease outcomes. Several systemic factors may contribute to this trend including Healthcare Access Gaps; Limited availability of diagnostic services, medication stock-outs and insufficient trained personnel in endemic regions. Operational Delays; Delayed reporting of test results, particularly for patients tested in the forenoon leads to missed treatment opportunities due to patient attrition before result collection. Socioeconomic Barriers; Work-related constraints and lack of awareness among patients and caregivers hinder timely treatment uptake.

Targeted interventions are warranted to address the rising trend of untreated malaria cases. These may include strengthen the point-of-care diagnostics and same-day result delivery. Enhancing community-based follow-up mechanisms. Prioritize resource allocation for *Pv* and *Pf* case management. Implement patient-centered strategies to improve treatment adherence.

### Analysis of Treated vs. Untreated Malaria Cases

The line graph illustrates the longitudinal trends in total treated malaria cases and the subset of patients who did not receive medication between 2002 and 2022, depicted in Figure 9. The number of treated cases peaked in 2005, reaches approximately 8,088, followed by a gradual decline over the subsequent years. A modest resurgence was observed in 2014, with 6,728 cases treated, but the overall trajectory remained downward, culminate in near-zero treated cases by 2022.

**Figure 9:**
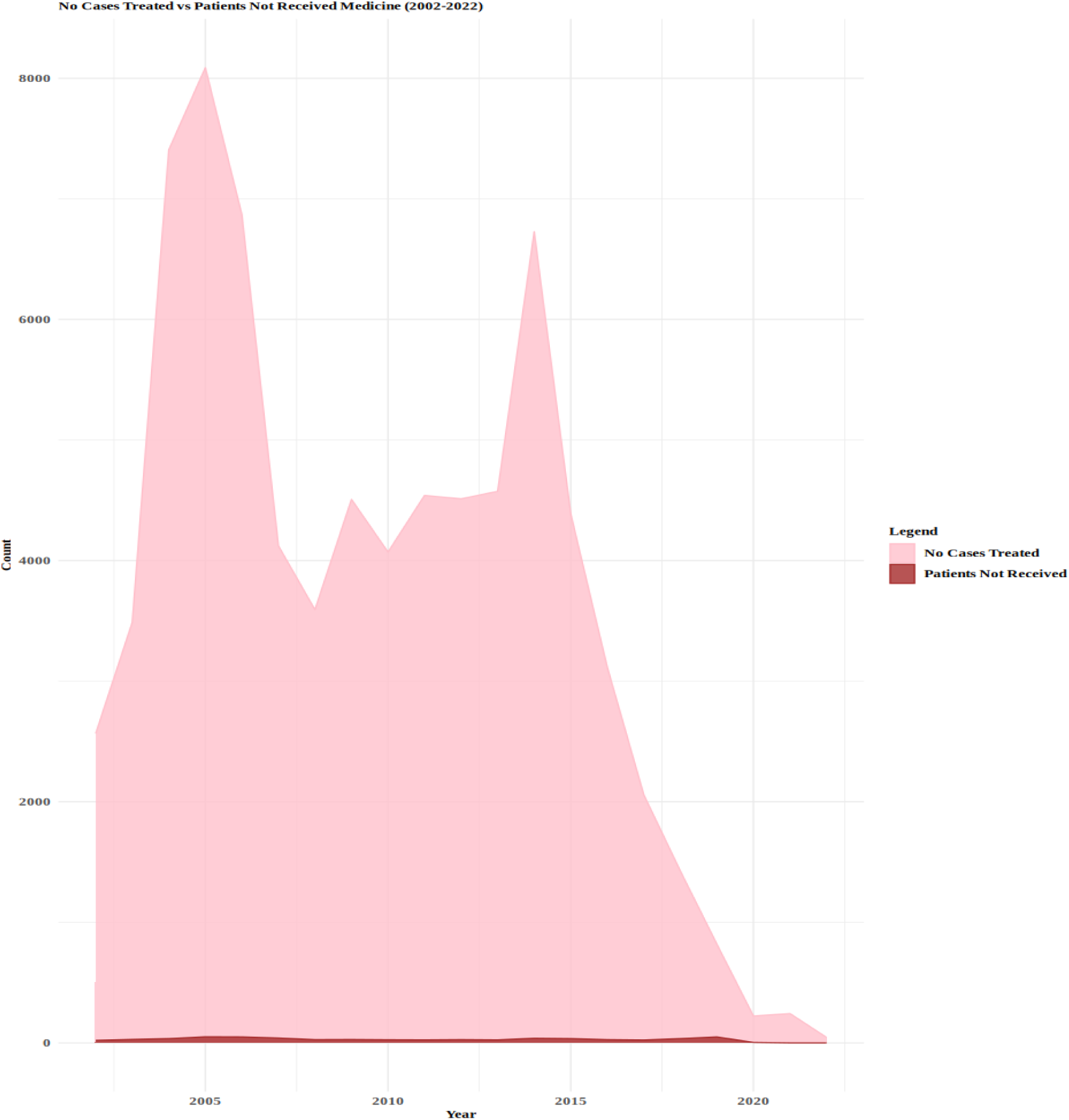
The graph covers that 21-year of malaria cases treated and patients not received medicine from 2002 to 2022. The year represents on the X-axis from 2002 to 2022. Number of cases represents on the Y-axis, ranging from 0 to 8000. Colour light pink area represents number of cases treated and darker pink area represents number of patients not received medicine.

The trend in patients not receive medication closely paralleled the treated case curve, with a peak around 2005 and a consistent decline thereafter. Notably, the gap between treated and untreated cases remained proportionally stable across the two decades, suggests persistent systemic challenges in ensuring complete medication delivery. In 2005, despite a treatment coverage rate of 99.22%, approximately 602 patients (0.77%) did not receive the prescribed antimalarial medication.

This discrepancy highlights critical gaps in healthcare delivery, particularly in medication accessibility and adherence. The consistently low proportion of untreated cases in later years reflects improvements in healthcare infrastructure and supply chain management. However, the presence of untreated cases, even during periods of high treatment coverage, underscores the need for targeted interventions to address logistical, socioeconomic and operational barriers in malaria case management.

### Follow-up Analysis of Treated Malaria Cases vs. Not Follow-up Cases

The annual proportions of treated malaria cases that received follow-up care versus those that did not, spanning the period from 2002 to 2022 are illustrated in Figure 10. Follow-up care plays a critical role in evaluate treatment efficacy, detect relapses and ensuring complete patient recovery. The data reveal four distinct phases. Initial Decline (2002–2006); Follow-up rates dropped from 24.16% in 2002 to 10.27% in 2006, indicating early challenges in sustaining post-treatment monitoring. Stable Low Rates (2006–2015); Follow-up care remained consistently low, ranging from 7.43% to 13.67%, while the proportion of cases without follow-up exceeded 85%, peaking at 90.91% in 2010. Improvement Phase (2017– 2019); A marked increase in follow-up care was observed, reaching 31.86% in 2019, the highest in the dataset suggests temporary improvements in healthcare delivery or programmatic support. Sharp Decline (2021); Follow-up rates fell drastically to 4.10%, while cases without follow-up rose to 95.90%, reflecting possible disruptions in healthcare services or reduced patient compliance.

**Figure 10:**
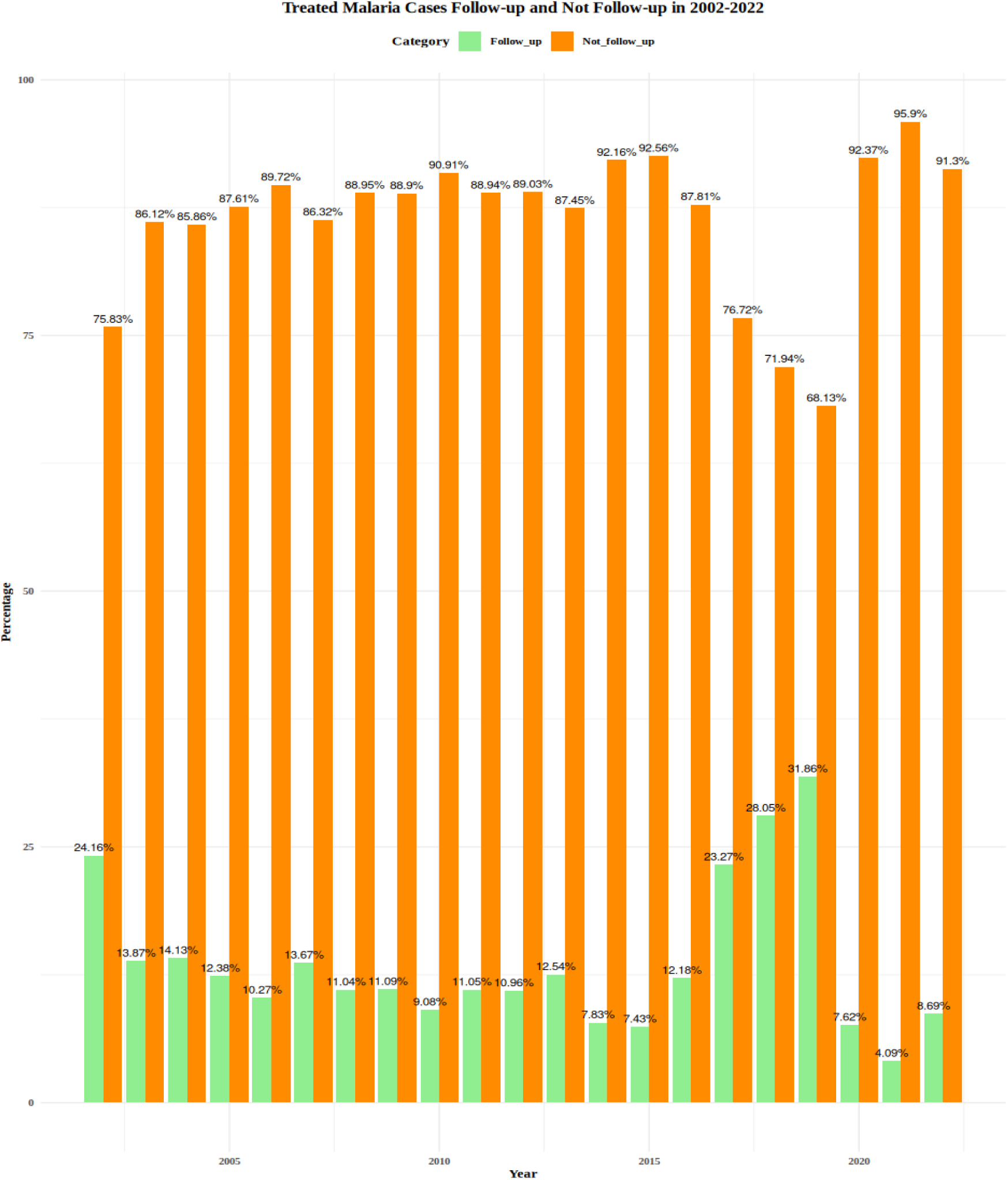
The graph shows a stacked bar chart displayed the percentage of treated malaria cases follow-up and don’t follow-up cases for each year from 2002 to 2022. The X-axis represents in year from 2002 to 2022 and Y-axis represents in percentage ranging from 0 to 100. The bar chart presents annual percentages of follow-up (green) and don’t follow-up (orange) cases, highlighting trends over a twenty-one-year period.

These trends underscore persistent gaps in post-treatment follow-up, likely driven by healthcare system constraints, patient-level barriers and variability in public health program implementation. Strengthen the follow-up mechanisms remains essential for effective malaria case management and long-term disease control.

## Discussion

Malaria elimination in India remains a formidable public health challenge, compounded by systemic gaps in treatment adherence, diagnostic precision and patient retention. These operational shortcomings, namely incomplete antimalarial medication courses, missed follow-ups, inpatient escape during treatment and lack of medication post-diagnosis are intricately linked to the emergence of antimalarial resistance, recurrent infections and treatment failures.

Incomplete antimalarial therapy is a critical driver of drug resistance^22^. Subtherapeutic exposure allows *Plasmodium* strains to survive and adapt^23^, leading to mutations in key resistance-associated genes such as *Pfcrt*, *Pfmdr1* and *K13*^24^. These mutations have been documented across endemic regions and are associated with resistance to artemisinin and partner drugs^24–25^. The persistence of resistant strains undermines mass drug administration (MDA) programs and necessitates the development of novel therapeutics ^25^. Follow-up on day 7 post-treatment is essential for detecting early treatment failure (ETF) and late treatment failure (LTF)^26–28^. However, socioeconomic constraints, particularly among migrant labourer, impede compliance. Studies have shown that over 87.51% of patients fail to return for follow-up, often due to wage loss or lack of employer support^29^. This gap contributes to undetected parasitaemia and continued transmission^30^.

Mixed infections involving *P. falciparum*, *P. vivax* and occasionally *P. malariae* or *P. ovale* are increasingly reported in India^31–32^. These co-infections complicate diagnosis and treatment, as misclassification can lead to inappropriate therapy and heightened resistance risk^33^. Molecular diagnostics such as nested polymerase chain reaction (nested PCR) and quantitative polymerase chain reaction (qPCR) offer improved sensitivity and specificity but remain underutilized due to cost and infrastructure limitations^34^. Notably, mixed infections are associated with severe clinical outcomes. Hypertension prevalence among mixed-infection patients reaches 98.32%, significantly higher than in mono-infections *P. falciparum* patients at 75.54% and *P. vivax* patients at 25.16%. This underscores the need for tailored therapeutic strategies and enhanced diagnostic capacity.

The COVID-19 pandemic and associated lockdowns led to a temporary decline in malaria cases, particularly in urban centres like Mangalore. The reverse migration of labourers typically at high risk due to poor housing and occupational exposure reduced local transmission. This phenomenon highlights the interplay between human mobility and malaria epidemiology as also observed in other regions^35^.

Malaria transmission in India exhibits pronounced seasonality, peaking during the monsoon months (June–September). Rainfall patterns directly influence vector breeding and parasite development^36–37^. Hydrological models have demonstrated that intraseasonal rainfall variability accounts for up to 39% of mosquito abundance variance^36^. Consequently, interventions such as indoor residual spraying (IRS) and insecticide-treated net (ITN) distribution should be timed to precede peak transmission periods^38^. Untreated malaria imposes significant economic strain on households through direct medical costs and productivity loss^39^. In endemic regions, prolonged illness due to incomplete treatment exacerbates poverty cycles and overwhelms healthcare systems^25,39^. Strengthening community health programs and ensuring drug availability are pivotal to break this cycle.

Policy Implications and Strategic Recommendations: Enhanced surveillance through the integration of molecular diagnostic tools into routine malaria monitoring systems is essential for detecting mixed infections and submicroscopic parasitaemia^34, 32^. To improve treatment adherence, community-based follow-up mechanisms should be implemented, supported by incentives that encourage patient retention and compliance with treatment protocols. Targeted interventions, such as indoor residual spraying (IRS) and insecticide-treated net (ITN) distribution campaigns, should be strategically aligned with seasonal transmission peaks using climate-informed predictive models ^36, 38^. For drug resistance monitoring, sentinel surveillance sites must be established to track resistance-associated genetic markers and conduct regular therapeutic efficacy studies^24–25^. To support socioeconomic needs and enhance treatment compliance among labourers, wage compensation schemes or flexible follow-up schedules should be introduced to address economic barriers to healthcare access.

## Conclusion

The 21-year longitudinal study provides a comprehensive overview of malaria epidemiology and case management in Mangalore, India, offering critical insights for national malaria elimination strategies. The findings reveal a substantial decline in *Plasmodium falciparum* infections from 45.15% in 2005 to 10.81% in 2016 indicating substantial progress in control measures. Conversely, *Plasmodium vivax* cases increased markedly, rising from 53.39% in 2005 to 86.95% in 2022, underscoring a shift in species dominance and the emerging threat posed by *P. vivax* in the region. Despite the overall reduction in malaria incidence, the gradual rise in mixed infections presents diagnostic and therapeutic challenges. These co-infections are often misclassified, increasing the risk of antimalarial resistance and complicating case management. The study also highlights a critically low day 7 follow-up rate (12.48%) among treated patients highlights critical gaps in post-treatment surveillance, which is essential for detect early treatment failure (ETF), late treatment failure (LTF), preventing relapse and curbing resistance development. Socioeconomic barriers including migration, wage loss, lack of employer support and limited healthcare access significantly affect treatment adherence and follow-up compliance. These barriers are especially pronounced among migrant labourers and daily wage workers, who often discontinue treatment prematurely or self-medicate without completing the full therapeutic course

To accelerate malaria elimination, targeted strategies are essential. Strengthen follow-up mechanisms, deploy mobile health interventions and community-based health workers can improve post-treatment monitoring. Diagnostic precision should be enhanced by incorporating molecular diagnostic such as nested PCR and qPCR, which can detect mixed infections and low-density parasitaemia. Expand access to healthcare outreach for vulnerable populations is crucial to improve availability of antimalarial drugs and diagnostic services. Public education and awareness campaigns should be promoted to support treatment adherence. Sustained vector control measures are needed including intensified distribution of long-lasting insecticidal nets (LLINs) and indoor residual spraying (IRS), especially in urban and peri-urban hotspots during seasonal transmission peaks. Regular monitoring of drug resistance is necessary including establish routine surveillance for resistance markers and advance the development of novel antimalarial therapies.

## Authors Detail’s

^1^ICMR-National Institute of Malaria Research, New Delhi-110077, India; Dr. Benudhar Mukhi, Mobile: +91-8197119414, ^2^Department of Biotechnology, Institute of Applied Sciences and Humanities, GLA University, Mathura, India; Mr. Chandan Kumar, ^3^ICMR-Regional Medical Research Centre, Bhubaneswar-751023, India; Dr. Mahesh Chandra Sahu. ^1^ICMR-National Institute of Malaria Research, New Delhi-110077, India; Prince Kumar, ^4^Department of Applied Zoology, Mangaluru University, Mangalore, India; Pradeep D. ^1^ICMR-National Institute of Malaria Research, New Delhi-110077, India; Dr. Anupkumar R. Anvikar. ^5^ICMR-National Institute of Malaria Research, ICMR Complex, Poojanhalli, Kannamangala Post, Devanhalli, Bangalore - 562110, India; Professor (Dr.) Susanta Kumar Ghosh, Telephone: +91-9845054366.

## Contributors

BM participated in collected clinical and epidemiological data and analysis the clinical data. AA, CK, MCS, PD, PK participated in data interpretation and critically reviewed this article. BM wrote this manuscript and all authors reviewed and approved the final manuscript.

## Data Sharing

The data used in this study are archived with Dr. Benudhar Mukhi and available from them upon reasonable request.

## Data Availability

All data produced in the present study are available upon reasonable request to the authors

## Acknowledgments

The authors acknowledge to Mr. Krishnaswamy, Senior Technician, Malaria Clinic, Dr. Rajeshwari Devi (District Surgeon and Superintendent), Wenlock District Government Hospital, Mangalore, Karnataka, India and Mr. M. Chandrasekhar Shenoy, Native volunteer from Mangalore, Karnataka, India for their support to this clinical malaria epidemiological study.

